# Changes in injecting versus smoking heroin, fentanyl, and methamphetamine among people who inject drugs in San Diego, California, 2020 to 2023

**DOI:** 10.1101/2024.02.23.24303293

**Authors:** William H. Eger, Daniela Abramovitz, Angela R. Bazzi, Annick Bórquez, Carlos F. Vera, Alicia Harvey-Vera, Joseph R. Friedman, Steffanie A. Strathdee

## Abstract

**Background:** Amidst a rapidly evolving drug supply in North America, people who inject drugs may be transitioning to smoking them. We aimed to assess changes in injecting and smoking heroin, fentanyl and methamphetamine among a cohort of people who injected drugs at baseline from San Diego, California.

**Methods:** Over five six-month periods spanning October 2020–April 2023, we assessed prevalence of injecting and smoking opioids or methamphetamine and whether participants used these drugs more frequently by smoking than injecting. Multivariable Poisson regression via Generalized Estimating Equations was used to examine time trends.

**Results:** Of 362 participants, median age was 40 years; most were male (72%), non-Hispanic (55%), and unhoused (67%). Among this cohort, of whom 100% injected (or injected and smoked) at baseline, by period five (two years later), 34% reported only smoking, while 59% injected and smoked, and 7% only injected. By period five, the adjusted relative risk (aRR) of injecting opioids was 0.41 (95% Confidence Interval [CI]: 0.33, 0.51) compared to period one, and the aRR for injecting methamphetamine was 0.50 (95% CI: 0.39, 0.63). Compared to period one, risks for smoking fentanyl rose significantly during period three (aRR=1.44, 95% CI: 1.06, 1.94), four (aRR=1.65, 95% CI: 1.24, 2.20) and five (aRR=1.90, 95% CI: 1.43, 2.53). Risks for smoking heroin and methamphetamine more frequently than injecting these drugs increased across all periods.

**Conclusions:** Opioid and methamphetamine injection declined precipitously, with notable increases in smoking these drugs. Research is urgently needed to understand the health consequences of these trends.

## 1. Introduction

The prevalence of injection drug use in the United States (U.S.) reportedly increased over the past decade in the context of multiple “waves” of the opioid crisis (1–3). With widespread contamination and replacement of heroin with illicitly manufactured fentanyls, drug-related overdose deaths surged between 2013 and 2021, and infectious disease transmission attributed to injection drug use has risen nationally (4–8). In addition to the unprecedented public health consequences of opioid use and injection, rising polysubstance use involving opioids and stimulants has presented additional health challenges (7, 9–12). In California, where fentanyl and methamphetamine are increasingly used in combination, the rate of fatal overdose has doubled since 2018 (7, 13).

Emerging evidence on the West Coast of the U.S. suggests that many people who previously injected black tar heroin have transitioned to smoking fentanyl (14, 15). For example, in San Francisco, California, one study found a significant decline in injecting heroin with a simultaneous increase in daily fentanyl smoking among people using opioids between 2018 and 2020 (14). In addition to individual preferences, characteristics of the drug supply can play a critical role in one’s decision to inject or smoke, indicating that the rapidly evolving drug supply across North America may be partially responsible for these emerging trends (15–17).

The extent to which transitioning from injecting to smoking drugs is occurring in the context of polysubstance use involving different types of unregulated opioids and stimulants is unknown because previous studies have not differentiated between heroin and fentanyl or assessed potential changes in the use of common stimulants such as methamphetamine. Understanding trends in consumption patterns of these specific drugs is vitally important in the context of the near replacement of heroin with fentanyl and in regions where overdose deaths increasingly involve both opioids and stimulants. To inform improved research and public health programmatic efforts, we assessed trends in injecting and smoking heroin, fentanyl, and methamphetamine in a cohort of people who inject drugs in San Diego, California, where the local drug supply has rapidly evolved in the context of ongoing cross-border drug trafficking and regional patterns of polysubstance use with opioids and methamphetamine (18, 19).

## 2. Materials and Methods

### 2.1. Participants and Eligibility

Participants were from the *La Frontera* cohort study, an ongoing, prospective investigation of HIV, Hepatitis C and drug overdose outcomes in the context of binational drug markets and cross-border mobility between San Diego, United States, and Tijuana, Mexico (20). Eligible participants were aged ≥18 years, reported past-month injection drug use (as evidenced through injection stigmata) and were living in San Diego County (hereafter: San Diego) or Tijuana at the time of screening. Half of the San Diego sample was intentionally recruited to reflect those who had crossed the border to use drugs in Tijuana within the past two years. Due to regional differences in fentanyl penetration and access to harm reduction services that may impact trends in injecting and smoking heroin, fentanyl, and methamphetamine (21, 22), this analysis was limited to participants who resided in San Diego who provided data between October 28, 2020, and April 27, 2023.

### 2.2. Data Collection

Trained bilingual interviewers collected data via a mobile outreach van that frequented locations with a high prevalence of drug use. Baseline recruitment occurred in two waves, October 28, 2020, to October 25, 2021 (Cohort 1; n=254) and February 7, 2022, to June 9, 2022 (Cohort 2; n=108). Following eligibility screening, participants provided written informed consent and completed interviewer-administered structured surveys at baseline, six-, 12-, 18-, and 24-months (see Supplemental Table 1). Participants received $20 USD compensation for each assessment. All study activities were approved by Institutional Review Boards at the University of California San Diego and Universidad Xochicalco in Tijuana.

### 2.3. Survey Measures

At each semi-annual visit, structured interviews gathered detailed data on sociodemographic characteristics, substance use behaviors and experiences of overdose, as described below.

*Outcomes of interest* included 1) prevalence of injecting and smoking heroin, fentanyl, or methamphetamine in the past six months; and 2) whether participants used these drugs more frequently by smoking than injecting. Each survey asked: 1) “During the last six months, on average, how often did you inject [drug]?” and 2) “During the last six months, on average, how often did you smoke, inhale, snort, or vape [drug]?” Responses included: never; one time per month or less; 2-3 days per month; one time per week; 2-3 days per week; 4-6 days per week; one time per day; 2-3 times per day; and ≥4 times per day. Participants could respond affirmatively to using any of the three drugs alone or in combination.

We first examined whether participants who used at least one of the three drugs (heroin, fentanyl, or methamphetamine) only injected, smoked or injected and only smoked any of the three drugs in the past six months by study period to determine the prevalence of injecting and smoking any drug over time. We then created eight binary variables corresponding to 1) whether a participant injected or smoked heroin, fentanyl, opioids (heroin and/or fentanyl use), or methamphetamine in the past six months for each period. Heroin and fentanyl use were combined to assess overall patterns in opioid use due to the replacement of heroin with fentanyl in the U.S.-Mexico border region during our study period, which may have led to misclassification (23).

For our second outcome, we created binary variables that indicated, among participants who used heroin, fentanyl, and/or methamphetamine in the previous six months, 2) whether participants used the drug more frequently via smoking than injecting.

*Our primary predictor of interest* was time period. We divided the data chronologically, regardless of study visit, into five mutually exclusive and consecutive six-month periods: 1) November 2020 to April 2021, 2) May 2021 to October 2021, 3) November 2021 to April 2022, 4) May 2022 to October 2022, and 5) November 2022 to April 2023.

*Additional variables* included age in years at baseline, sex assigned at birth (female or male), ethnicity (non-Hispanic/Latino or Hispanic/Latino/Mexican), country of birth (U.S. or other), years of education completed, monthly income (≥500 USD or <500 USD), past six-month housing status (housed or unhoused), incarceration status (no or yes), receptive needle sharing (no or yes), overdose experience (no or yes) and use of methadone, buprenorphine or other drug treatment (no or yes). We also summarized years of injection drug use, current smoking of cigarettes (no or yes), and past six-month marijuana use (no or yes).

### 2.4. Statistical Analysis

We first described the sample according to baseline sociodemographic characteristics, substance use behaviors (e.g., past six-month injecting and smoking of heroin, fentanyl, and methamphetamine) and overdose experiences by calculating frequencies and percentages for categorical variables and medians and interquartile ranges for continuous variables. Frequencies and percentages were used to describe the distribution of use and whether participants used more frequently via smoking than injecting for each period. We also examined, among participants who used at least one of the three drugs, whether participants only injected, only smoked and injected and smoked (any of the three drugs) in the past six months by study period to determine the prevalence of injecting and smoking any drug over time.

Next, to assess differential attrition in baseline characteristics between participants who completed at least one follow-up visit and those who did not, we used Chi-Square, Fisher’s exact and Mann-Whitney tests (for binary and continuous variables, respectively). We also conducted a similar supplemental analysis with baseline characteristics to compare each recruitment wave, which revealed that compared to cohort two, participants in the first cohort were more likely to be Hispanic/Latino/Mexican, make less than 500 USD per month, and report past six-month smoking and injecting of heroin, receptive needle sharing, and methadone, buprenorphine, or other drug treatment enrollment (p<0.05; not reported). Due to these differences by cohort, we controlled for “recruitment wave” in multivariable models.

We then used multivariable Poisson regressions via generalized estimating equations with robust standard errors and an unstructured covariance matrix to assess potential changes in use of each drug over time (22, 24). Time period was the primary predictor, with later periods being compared to period one concerning the risk of using each drug by 1) injecting or smoking and by 2) smoking more often than injecting. All multivariable models were adjusted for recruitment wave, age, sex assigned at birth and past six-month use of methadone, buprenorphine, or other drug treatment, which were selected *a priori* based on the literature (14). Interpretations were based on the magnitude of the adjusted relative risk (aRR) and corresponding 95% confidence intervals (25, 26). All analyses were conducted using SAS version 9.4; graphs were made in R version 4.3.2.

## 3. Results

### 3.1. Sample Characteristics

The analytical sample consisted of 362 unique participants who completed ≥1 study visit during the study period, comprising 876 surveys and a median of two visits per participant (interquartile range [IQR]=2.0, 4.0). Most (77%) had more than one study visit before the cut-off date for this analysis. Approximately 6% (n=21) died before April 31, 2023, and did not contribute follow-up data. Those who did not have more than one study visit were younger (p=0.03), but no other significant differences emerged between the groups.

At baseline, median age was 40 years (IQR=33.0, 52.0; Table 1). A minority were born in another country (5%), assigned female at birth (29%) and were Hispanic/Latino/Mexican (45%). Two-thirds (67%) reported being unhoused and 15% were incarcerated in the past six months. At baseline, most injected heroin (72%) and methamphetamine (70%) and also smoked, snorted, inhaled or vaped methamphetamine (81%) in the past six months. The past six-month prevalence of receptive syringe sharing (34%) and overdose (20%) was high, while only 13% were enrolled in methadone, buprenorphine, or other drug treatment.

**Table 1.**
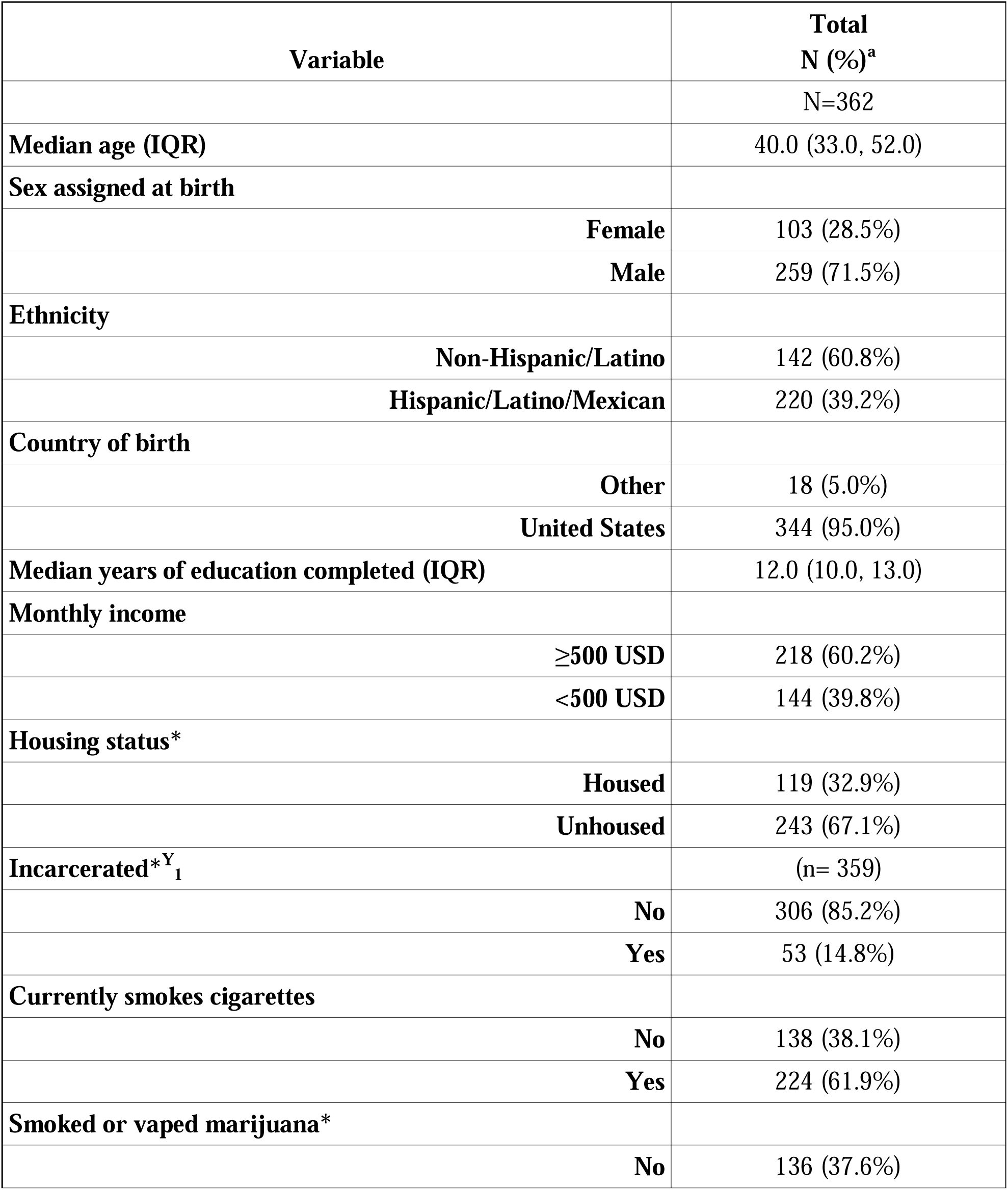
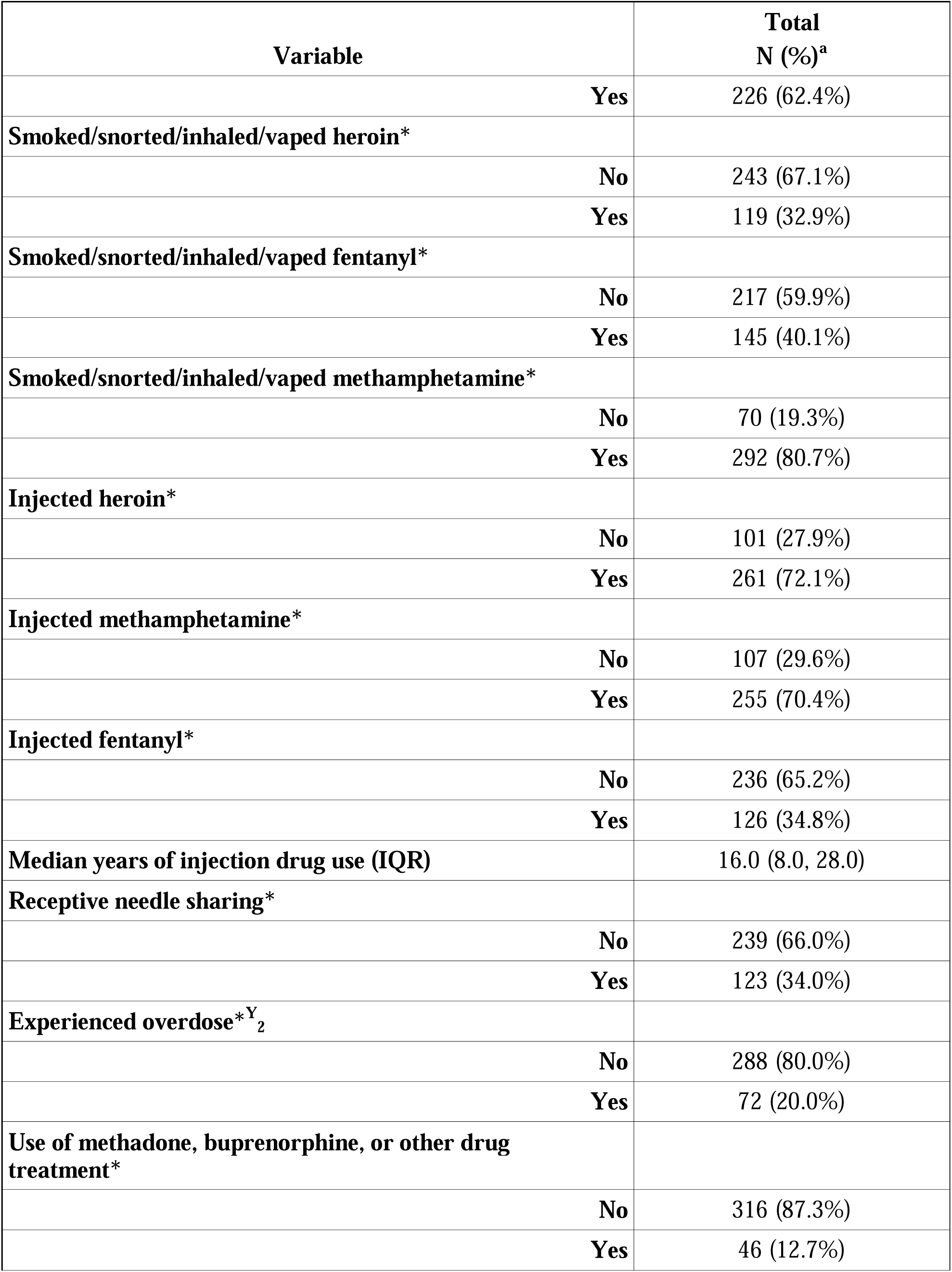
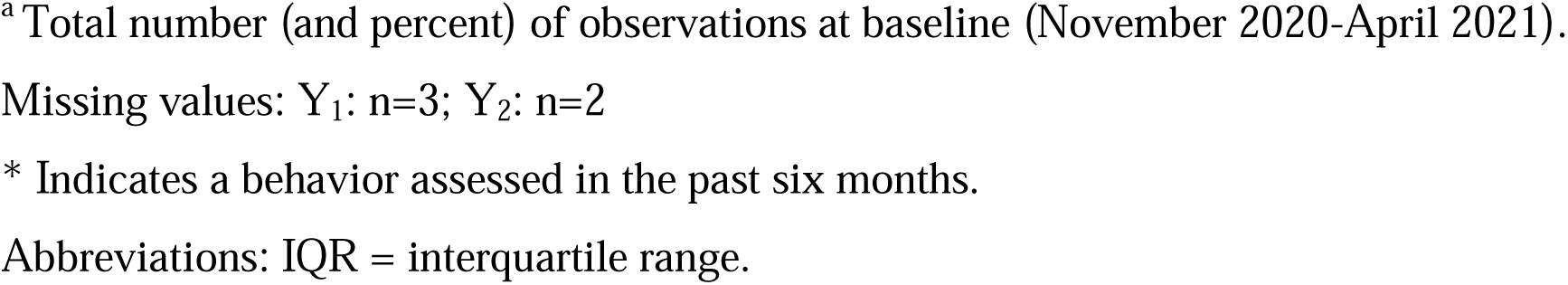
Baseline characteristics of study participants residing in San Diego County, California, between October 28, 2020, and October 25, 2021.

### 3.2. Prevalence of injecting only, smoking only, or injecting and smoking

As represented in Figure 1 (and Supplemental Table 2), the prevalence of only injecting decreased over time, dropping from 15.6% in period one to 6.5% in period five. Additionally, the prevalence of injecting and smoking decreased over time, falling from 84.4% of the sample in period one to 59.4% in period five. Conversely, the fraction of participants only smoking increased over time, with none reporting this behavior in period one (by definition, as only people who injected drugs were included in the study) and 34.2% reporting it in period five.

**Figure 1.**
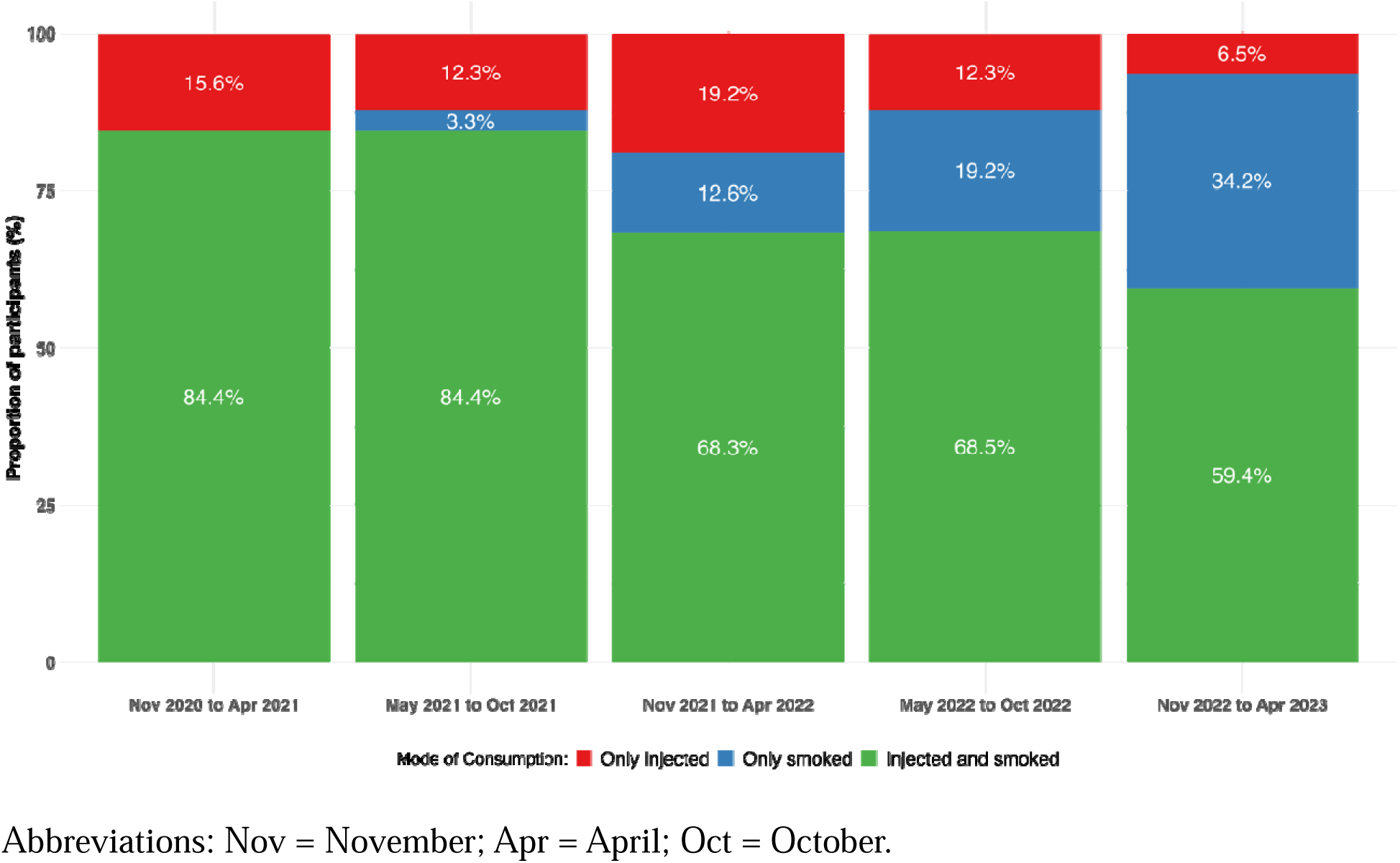
Prevalence of injecting only, smoking only, and injecting and smoking (heroin, fentanyl, or methamphetamine) in the past six months among study participants residing in San Diego County, California, who used heroin, fentanyl, or methamphetamine in the past six months between October 28, 2020, and April 31, 2023

### 3.3. Prevalence of injecting and smoking heroin, fentanyl, opioids, or methamphetamine among the total sample

As shown in Table 2 and corresponding Figure 2, the risk of using heroin decreased significantly in period five (vs. period one; aRR=0.28; 95% confidence interval [CI]: 0.21, 0.39). The risk of injecting heroin in the past six months also declined by 46% in period three (aRR=0.54; 95% CI: 0.44, 0.66), 54% in period four (aRR=0.46; 95% CI: 0.38, 0.57) and 76% in period five (aRR=0.24; 95% CI: 0.17, 0.34) compared to period one. Similarly, the risk of smoking heroin in the past six months declined in periods three (aRR=0.42; 95% CI: 0.27, 0.67), four (aRR=0.35; 95% CI: 0.23, 0.54), and five (aRR=0.30; 95% CI: 0.17, 0.50) relative to period one.

**Figure 2.**
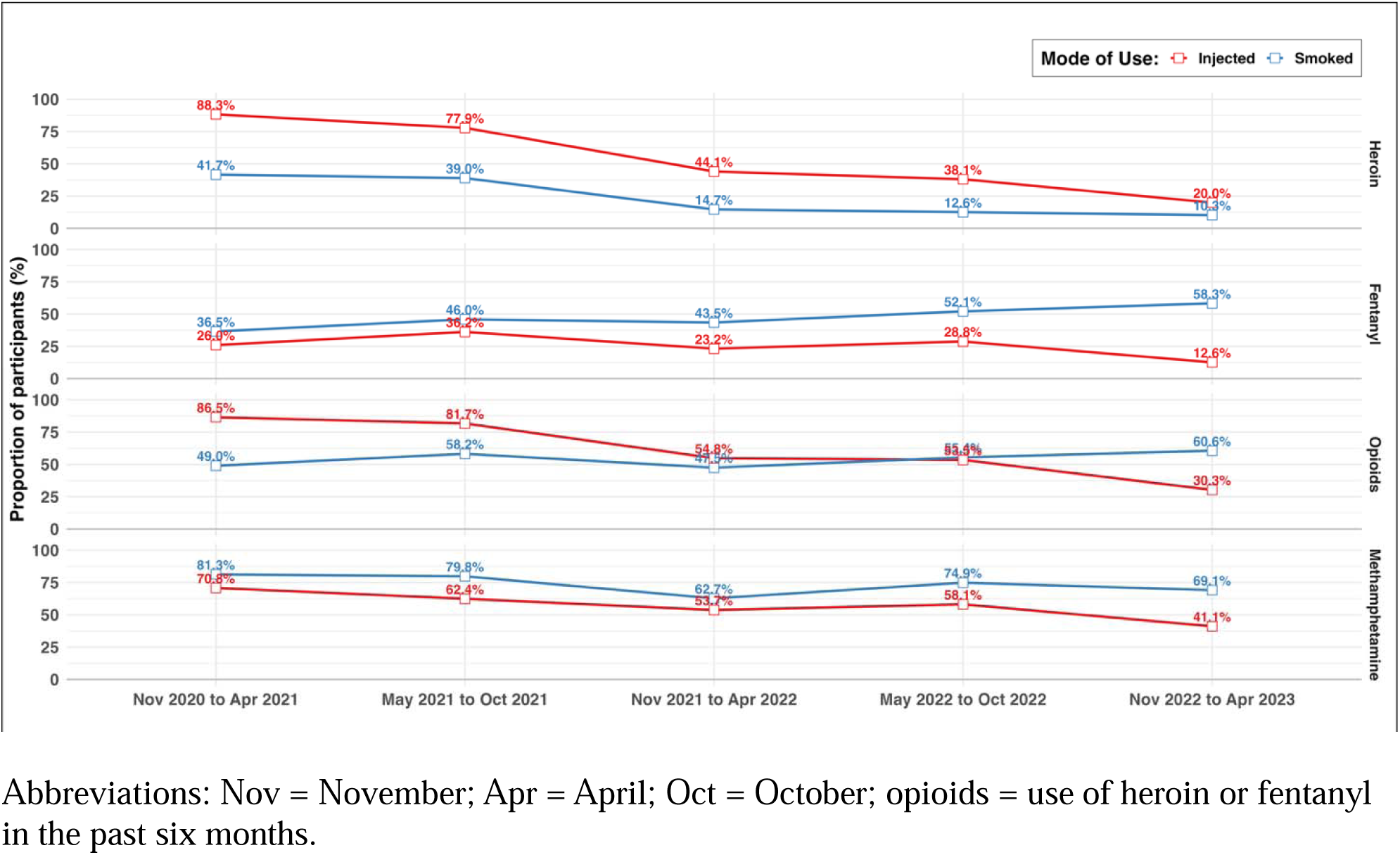
Prevalence of injecting or smoking heroin, fentanyl, opioids, or methamphetamine among study participants residing in San Diego County, California, between October 28, 2020, and April 31, 2023

**Table 2.**
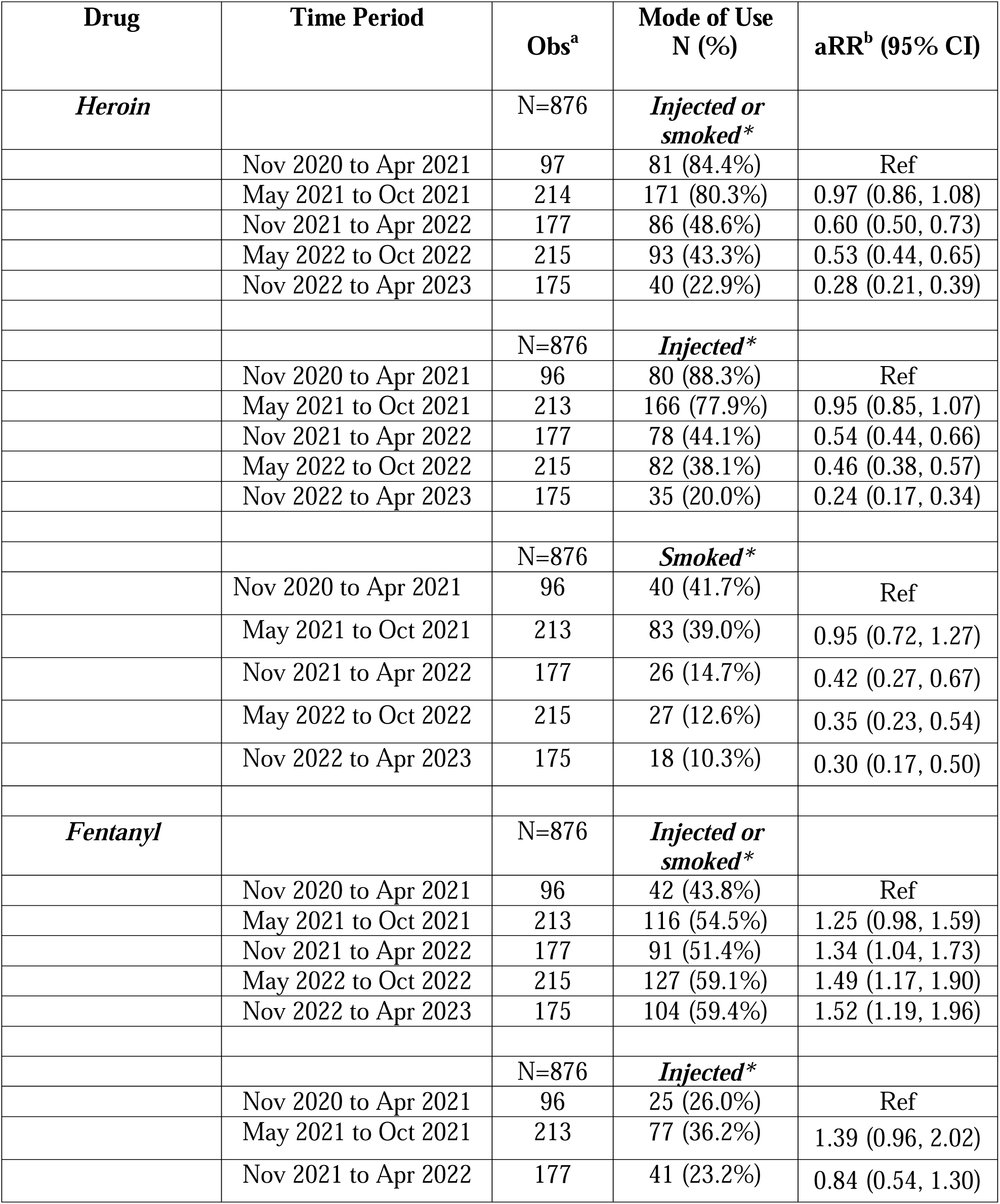
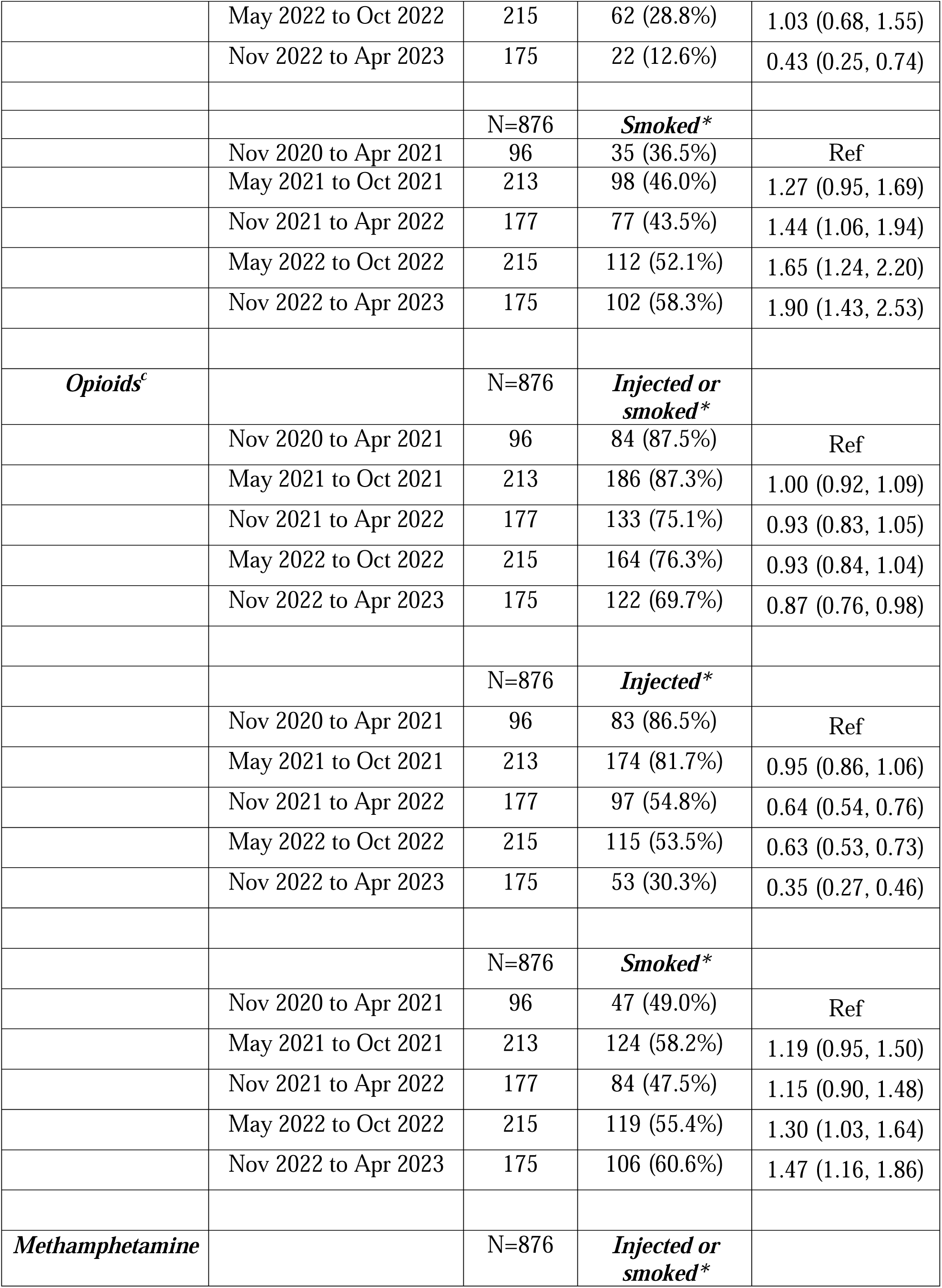
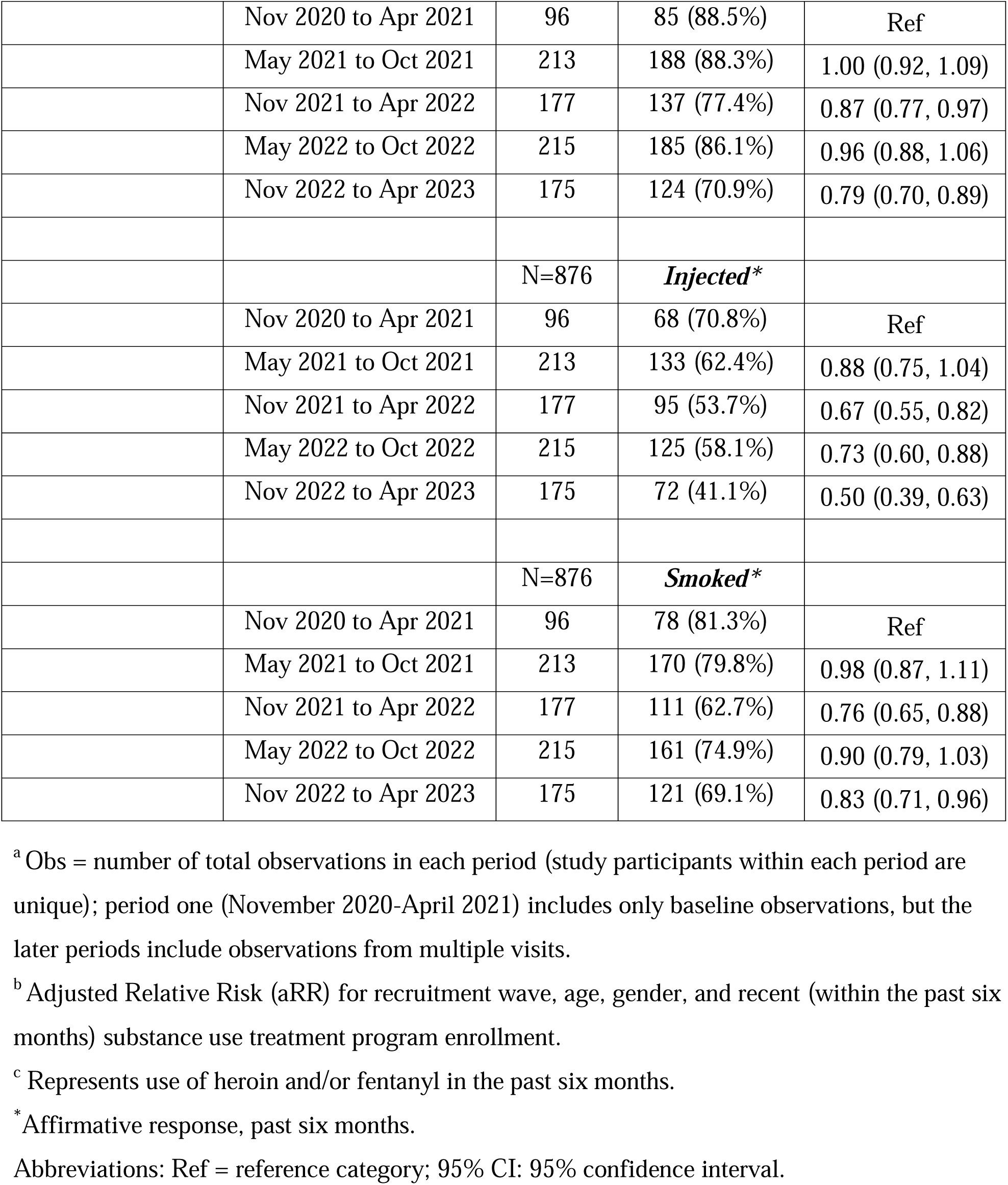
Prevalence of injecting and smoking heroin, fentanyl, or methamphetamine among study participants residing in San Diego County, California, between October 28, 2020, and April 31, 2023.

| Drug | Time Period | Obs<sup>a</sup> | Mode of Use<br>N (%) | aRR<sup>b</sup> (95% CI) |
| --- | --- | --- | --- | --- |
| *Heroin* |  | N=876 | *Injected or smoked\** |  |
|  | Nov 2020 to Apr 2021 | 97 | 81 (84.4%) | Ref |
|  | May 2021 to Oct 2021 | 214 | 171 (80.3%) | 0.97 (0.86, 1.08) |
|  | Nov 2021 to Apr 2022 | 177 | 86 (48.6%) | 0.60 (0.50, 0.73) |
|  | May 2022 to Oct 2022 | 215 | 93 (43.3%) | 0.53 (0.44, 0.65) |
|  | Nov 2022 to Apr 2023 | 175 | 40 (22.9%) | 0.28 (0.21, 0.39) |
|  |  | N=876 | *Injected\** |  |
|  | Nov 2020 to Apr 2021 | 96 | 80 (88.3%) | Ref |
|  | May 2021 to Oct 2021 | 213 | 166 (77.9%) | 0.95 (0.85, 1.07) |
|  | Nov 2021 to Apr 2022 | 177 | 78 (44.1%) | 0.54 (0.44, 0.66) |
|  | May 2022 to Oct 2022 | 215 | 82 (38.1%) | 0.46 (0.38, 0.57) |
|  | Nov 2022 to Apr 2023 | 175 | 35 (20.0%) | 0.24 (0.17, 0.34) |
|  |  | N=876 | *Smoked\** |  |
|  | Nov 2020 to Apr 2021 | 96 | 40 (41.7%) | Ref |
|  | May 2021 to Oct 2021 | 213 | 83 (39.0%) | 0.95 (0.72, 1.27) |
|  | Nov 2021 to Apr 2022 | 177 | 26 (14.7%) | 0.42 (0.27, 0.67) |
|  | May 2022 to Oct 2022 | 215 | 27 (12.6%) | 0.35 (0.23, 0.54) |
|  | Nov 2022 to Apr 2023 | 175 | 18 (10.3%) | 0.30 (0.17, 0.50) |
| *Fentanyl* |  | N=876 | *Injected or smoked\** |  |
|  | Nov 2020 to Apr 2021 | 96 | 42 (43.8%) | Ref |
|  | May 2021 to Oct 2021 | 213 | 116 (54.5%) | 1.25 (0.98, 1.59) |
|  | Nov 2021 to Apr 2022 | 177 | 91 (51.4%) | 1.34 (1.04, 1.73) |
|  | May 2022 to Oct 2022 | 215 | 127 (59.1%) | 1.49 (1.17, 1.90) |
|  | Nov 2022 to Apr 2023 | 175 | 104 (59.4%) | 1.52 (1.19, 1.96) |
|  |  | N=876 | *Injected\** |  |
|  | Nov 2020 to Apr 2021 | 96 | 25 (26.0%) | Ref |
|  | May 2021 to Oct 2021 | 213 | 77 (36.2%) | 1.39 (0.96, 2.02) |
|  | Nov 2021 to Apr 2022 | 177 | 41 (23.2%) | 0.84 (0.54, 1.30) |
|  | May 2022 to Oct 2022 | 215 | 62 (28.8%) | 1.03 (0.68, 1.55) |
|  | Nov 2022 to Apr 2023 | 175 | 22 (12.6%) | 0.43 (0.25, 0.74) |
|  |  | N=876 | ***Smoked\**** |  |
|  | Nov 2020 to Apr 2021 | 96 | 35 (36.5%) | Ref |
|  | May 2021 to Oct 2021 | 213 | 98 (46.0%) | 1.27 (0.95, 1.69) |
|  | Nov 2021 to Apr 2022 | 177 | 77 (43.5%) | 1.44 (1.06, 1.94) |
|  | May 2022 to Oct 2022 | 215 | 112 (52.1%) | 1.65 (1.24, 2.20) |
|  | Nov 2022 to Apr 2023 | 175 | 102 (58.3%) | 1.90 (1.43, 2.53) |
| ***Opioids<sup>c</sup>*** |  | N=876 | ***Injected or smoked\**** |  |
|  | Nov 2020 to Apr 2021 | 96 | 84 (87.5%) | Ref |
|  | May 2021 to Oct 2021 | 213 | 186 (87.3%) | 1.00 (0.92, 1.09) |
|  | Nov 2021 to Apr 2022 | 177 | 133 (75.1%) | 0.93 (0.83, 1.05) |
|  | May 2022 to Oct 2022 | 215 | 164 (76.3%) | 0.93 (0.84, 1.04) |
|  | Nov 2022 to Apr 2023 | 175 | 122 (69.7%) | 0.87 (0.76, 0.98) |
|  |  | N=876 | ***Injected\**** |  |
|  | Nov 2020 to Apr 2021 | 96 | 83 (86.5%) | Ref |
|  | May 2021 to Oct 2021 | 213 | 174 (81.7%) | 0.95 (0.86, 1.06) |
|  | Nov 2021 to Apr 2022 | 177 | 97 (54.8%) | 0.64 (0.54, 0.76) |
|  | May 2022 to Oct 2022 | 215 | 115 (53.5%) | 0.63 (0.53, 0.73) |
|  | Nov 2022 to Apr 2023 | 175 | 53 (30.3%) | 0.35 (0.27, 0.46) |
|  |  | N=876 | ***Smoked\**** |  |
|  | Nov 2020 to Apr 2021 | 96 | 47 (49.0%) | Ref |
|  | May 2021 to Oct 2021 | 213 | 124 (58.2%) | 1.19 (0.95, 1.50) |
|  | Nov 2021 to Apr 2022 | 177 | 84 (47.5%) | 1.15 (0.90, 1.48) |
|  | May 2022 to Oct 2022 | 215 | 119 (55.4%) | 1.30 (1.03, 1.64) |
|  | Nov 2022 to Apr 2023 | 175 | 106 (60.6%) | 1.47 (1.16, 1.86) |
| ***Methamphetamine*** |  | N=876 | ***Injected or smoked\**** |  |
|  | Nov 2020 to Apr 2021 | 96 | 85 (88.5%) | Ref |
|  | May 2021 to Oct 2021 | 213 | 188 (88.3%) | 1.00 (0.92, 1.09) |
|  | Nov 2021 to Apr 2022 | 177 | 137 (77.4%) | 0.87 (0.77, 0.97) |
|  | May 2022 to Oct 2022 | 215 | 185 (86.1%) | 0.96 (0.88, 1.06) |
|  | Nov 2022 to Apr 2023 | 175 | 124 (70.9%) | 0.79 (0.70, 0.89) |
|  |  | N=876 | ***Injected\**** |  |
|  | Nov 2020 to Apr 2021 | 96 | 68 (70.8%) | Ref |
|  | May 2021 to Oct 2021 | 213 | 133 (62.4%) | 0.88 (0.75, 1.04) |
|  | Nov 2021 to Apr 2022 | 177 | 95 (53.7%) | 0.67 (0.55, 0.82) |
|  | May 2022 to Oct 2022 | 215 | 125 (58.1%) | 0.73 (0.60, 0.88) |
|  | Nov 2022 to Apr 2023 | 175 | 72 (41.1%) | 0.50 (0.39, 0.63) |
|  |  | N=876 | ***Smoked\**** |  |
|  | Nov 2020 to Apr 2021 | 96 | 78 (81.3%) | Ref |
|  | May 2021 to Oct 2021 | 213 | 170 (79.8%) | 0.98 (0.87, 1.11) |
|  | Nov 2021 to Apr 2022 | 177 | 111 (62.7%) | 0.76 (0.65, 0.88) |
|  | May 2022 to Oct 2022 | 215 | 161 (74.9%) | 0.90 (0.79, 1.03) |
|  | Nov 2022 to Apr 2023 | 175 | 121 (69.1%) | 0.83 (0.71, 0.96) |

<sup>a</sup> Obs = number of total observations in each period (study participants within each period are unique); period one (November 2020-April 2021) includes only baseline observations, but the later periods include observations from multiple visits.
<sup>b</sup> Adjusted Relative Risk (aRR) for recruitment wave, age, gender, and recent (within the past six months) substance use treatment program enrollment.
<sup>c</sup> Represents use of heroin and/or fentanyl in the past six months.
\* Affirmative response, past six months.
Abbreviations: Ref = reference category; 95% CI: 95% confidence interval.

The risk of using fentanyl increased by 34% in period three (aRR=1.34; 95% CI: 1.04, 1.73), 49% in period four (aRR=1.49; 95% CI: 1.17, 1.90), and 52% in period five (aRR=1.52; 95% CI: 1.19, 1.96). The prevalence of injecting fentanyl was stable across periods one through four but was 57% lower in period five (vs. period one; aRR=0.43; 95% CI: 0.25, 0.74). In contrast, the risk of smoking fentanyl in the past six months increased by 44% in period three (aRR=1.44; 95% CI: 1.06, 1.94), 65% in period four (aRR=1.65; 95% CI: 1.24, 2.20), and 90% in period five (aRR=1.90; 95% CI: 1.43, 2.53) compared to period one.

The risk of using opioids was stable from periods one to four but decreased by 13% in period five (vs. period one; aRR=0.87; 95% CI: 0.76, 0.98). The risk of injecting opioids declined by 31% in period three (aRR=0.69; 95% CI: 61, 0.78), 33% in period four (aRR=0.67; 95% CI: 0.60, 0.76), and 59% in period five (aRR=0.41; 95% CI: 0.33, 0.51). The risk of smoking opioids, meanwhile, increased by 25% in period three (aRR=1.25; 95% CI: 1.00, 1.56), 39% in period four (aRR=1.39; 95% CI: 1.13, 1.72), and 69% in period five (aRR=1.69; 95% CI: 1.37, 2.07) relative to period one.

The risk of methamphetamine use also decreased over time, with a 21% decline in risk in period five (vs. period one; aRR=0.79; 95% CI: 0.70, 0.89). The risk of injecting and smoking methamphetamine also decreased, with a more marked decline in the risk of injecting than smoking. For instance, in period five, the risk of injecting methamphetamine decreased by 50% (vs. period one; aRR=0.50; 95% CI: 0.39, 0.63) while the risk of smoking methamphetamine decreased by 17% (vs. period one; aRR=0.83; 95% CI: 0.71, 0.96).

### 3.4. Smoking heroin, fentanyl, opioids, or methamphetamine more often than injecting

Among participants who used heroin (Table 3 and Figure 3), the risk of injecting it declined by 14% between periods one and five (aRR=0.86; 95% CI 0.77, 0.97). The prevalence of smoking heroin remained stable over time, with no significant changes from period one over the study period. However, relative to period one, the risk of smoking heroin more often than injecting it increased by 169% in period three (aRR=2.69; 95% CI: 1.11, 6.49), 158% in period four (aRR=2.58; 95% CI: 1.08, 6.16) and 249% in period five (aRR=3.49; 95% CI: 1.39, 8.74).

**Figure 3.**
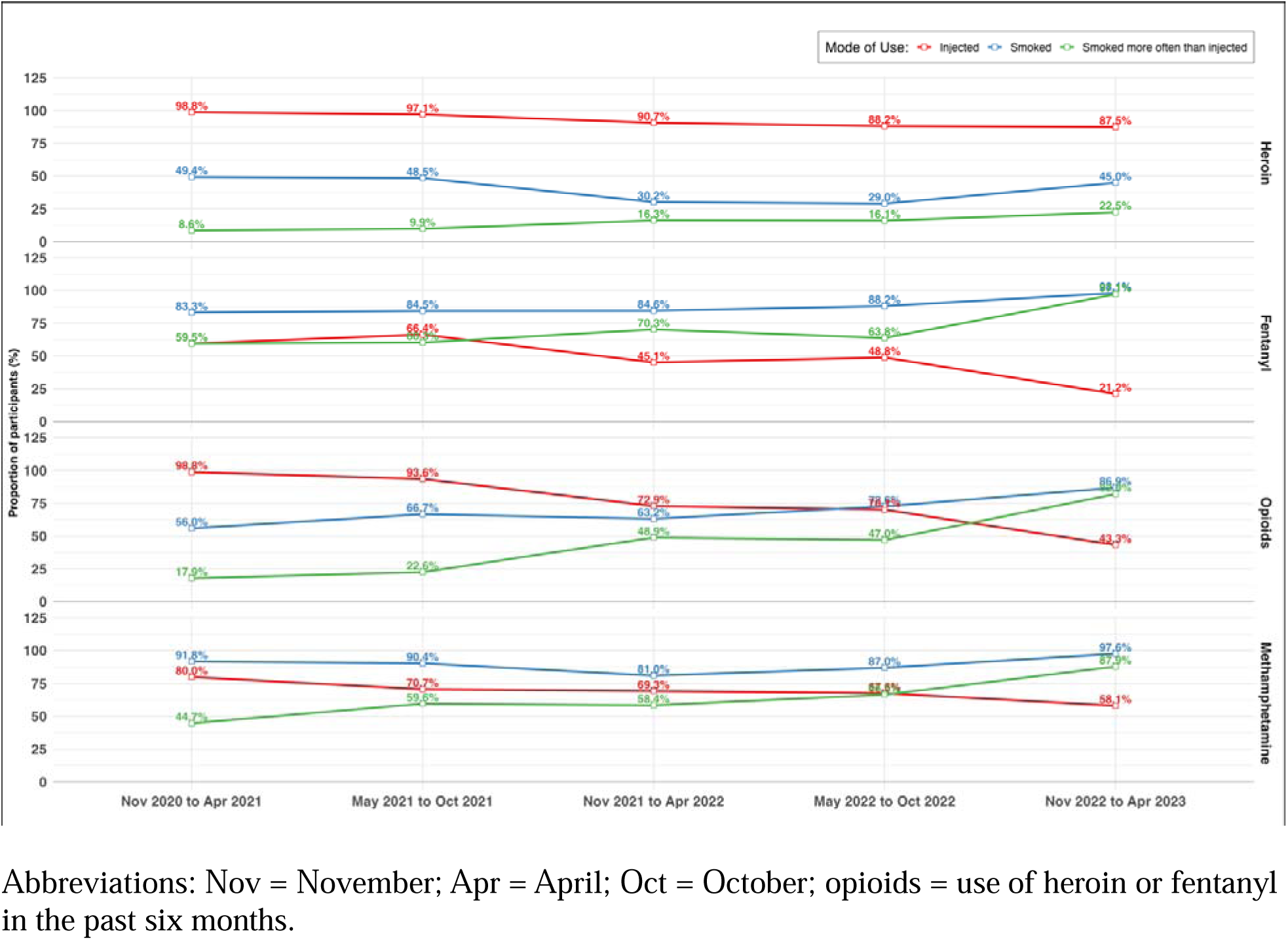
Prevalence of smoking heroin, fentanyl, opioids, or methamphetamine more often than injecting among study participants residing in San Diego County, California, who used heroin, fentanyl, or methamphetamine in the past six months, between October 28, 2020, and April 31, 2023

**Table 3.**
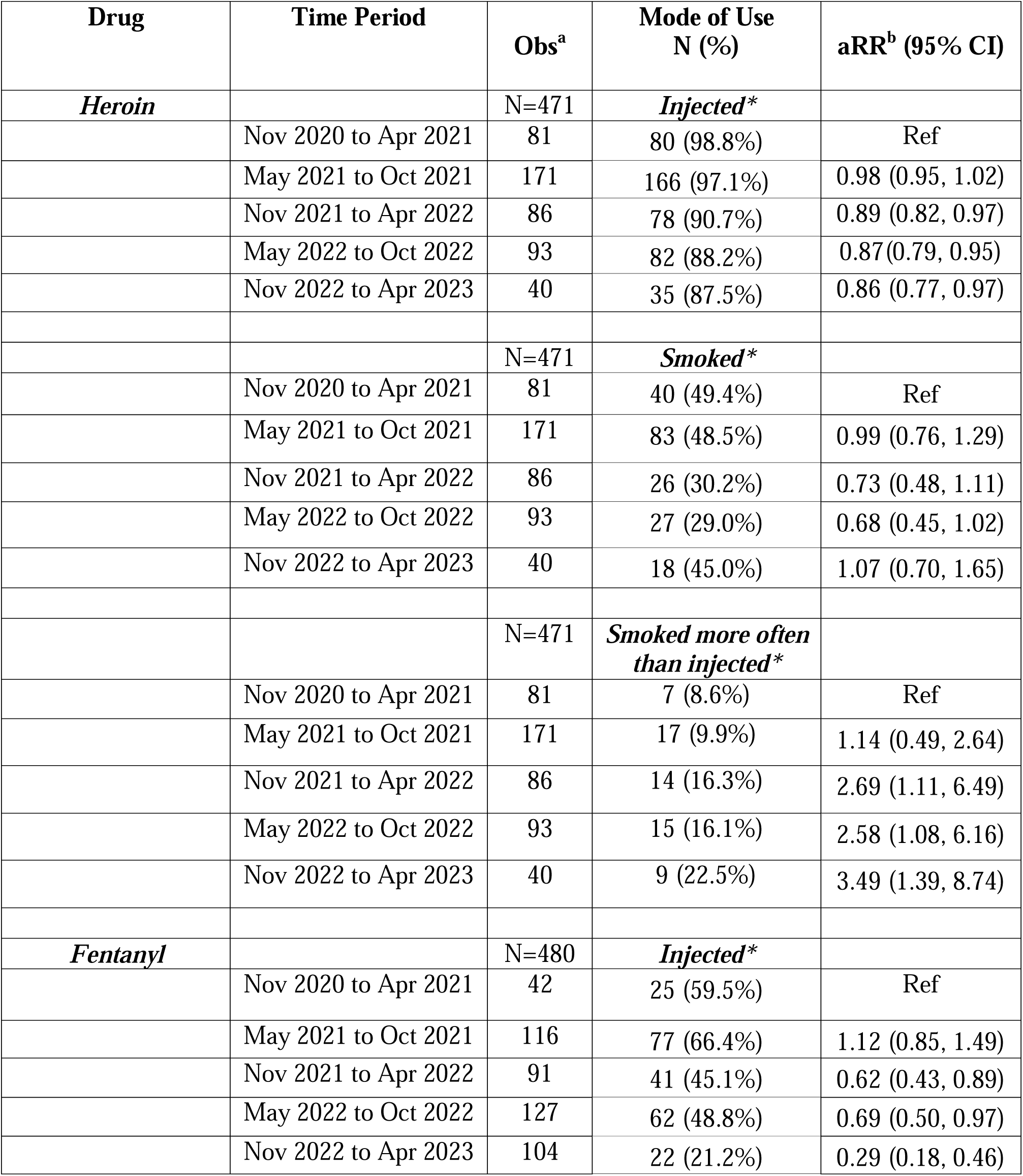
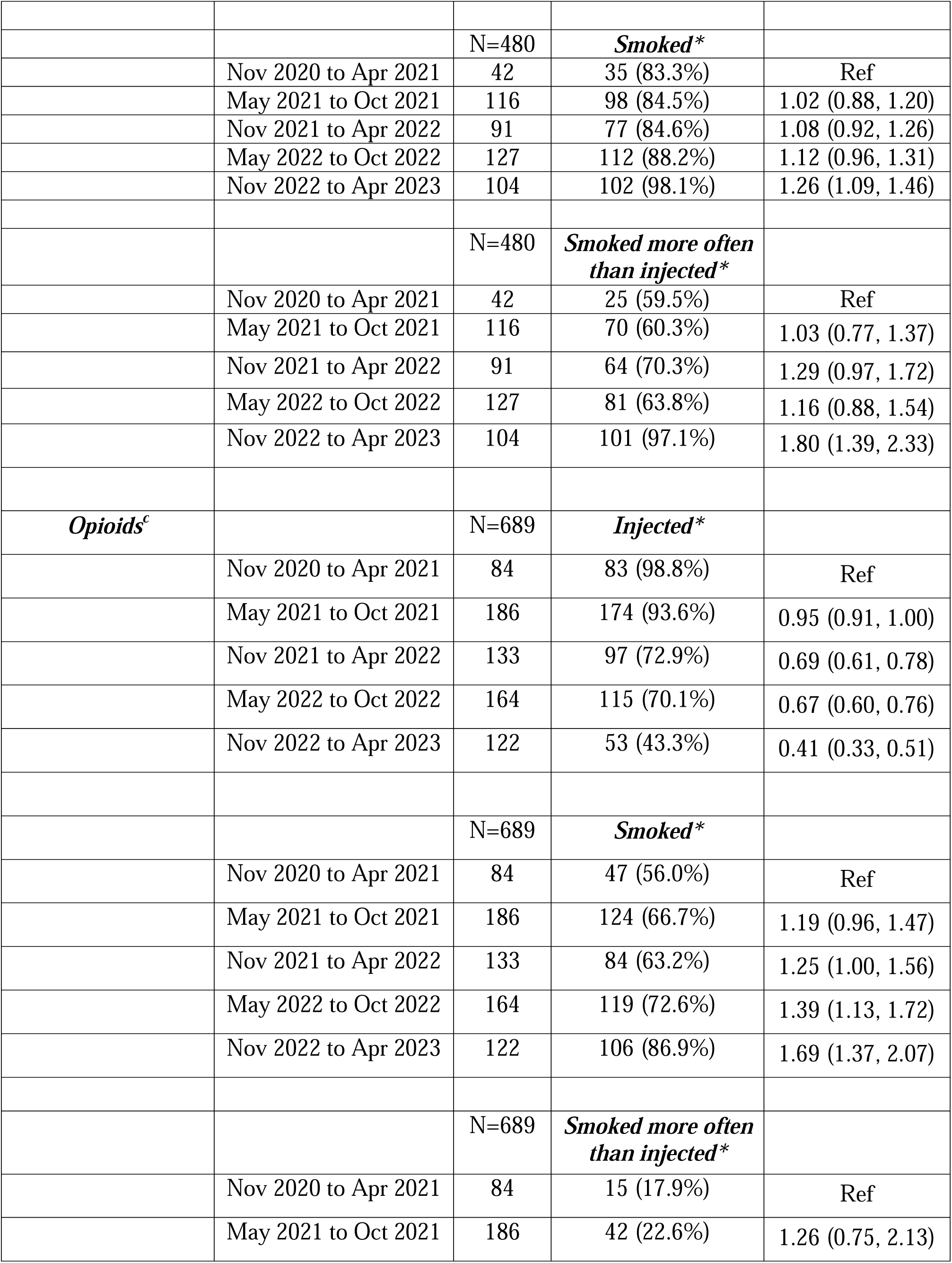
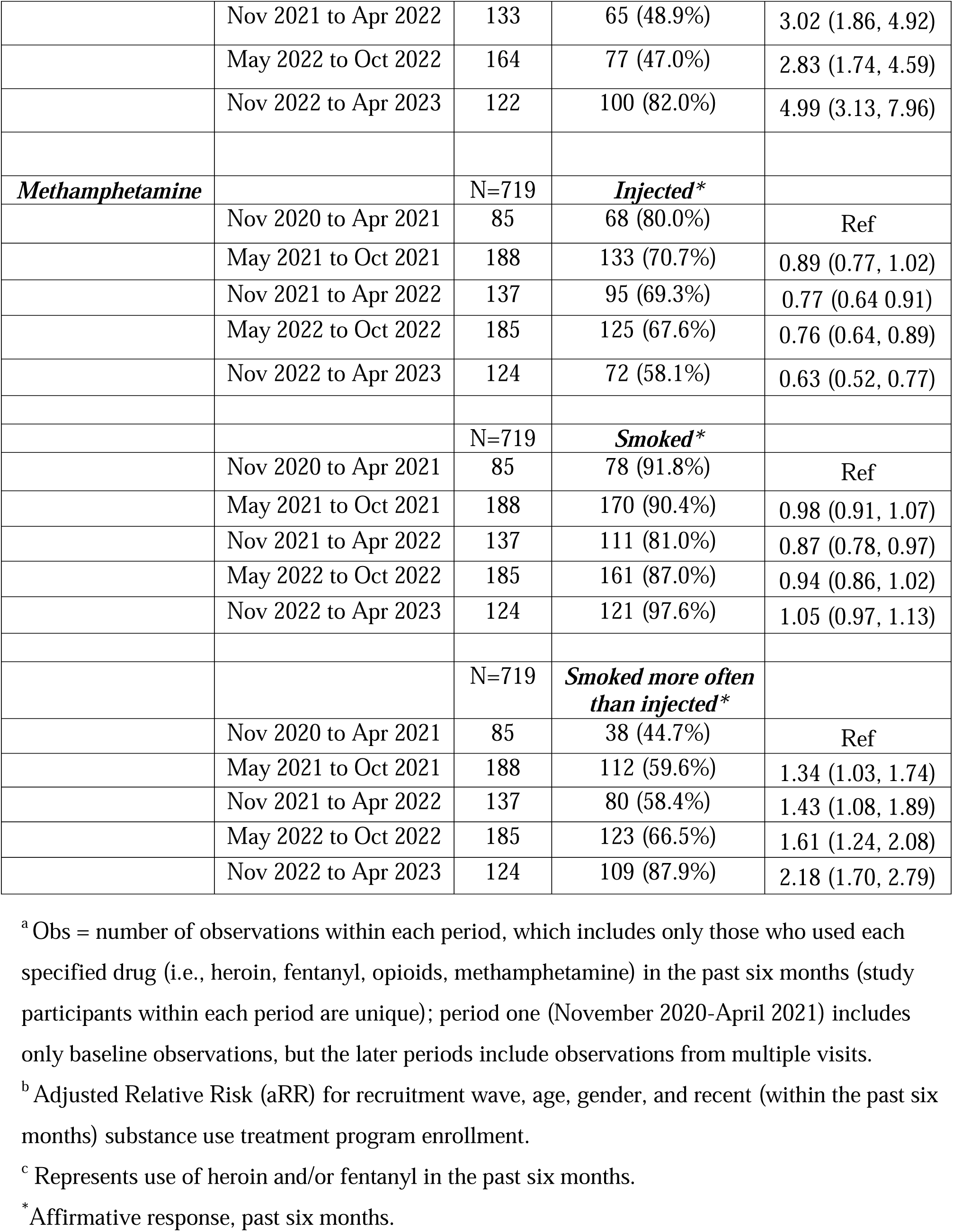
Prevalence of smoking heroin, fentanyl, opioids, or methamphetamine more often than injecting among study participants residing in San Diego County, California, who used heroin, fentanyl, opioids, or methamphetamine in the past six months, between October 28, 2020, and April 31, 2023.

| **Drug** | **Time Period** | **Obs<sup>a</sup>** | **Mode of Use**<br>**N (%)** | **aRR<sup>b</sup> (95% CI)** |
| --- | --- | --- | --- | --- |
| ***Heroin*** |  | N=471 | ***Injected\**** |  |
|  | Nov 2020 to Apr 2021 | 81 | 80 (98.8%) | Ref |
|  | May 2021 to Oct 2021 | 171 | 166 (97.1%) | 0.98 (0.95, 1.02) |
|  | Nov 2021 to Apr 2022 | 86 | 78 (90.7%) | 0.89 (0.82, 0.97) |
|  | May 2022 to Oct 2022 | 93 | 82 (88.2%) | 0.87(0.79, 0.95) |
|  | Nov 2022 to Apr 2023 | 40 | 35 (87.5%) | 0.86 (0.77, 0.97) |
|  |  | N=471 | ***Smoked\**** |  |
|  | Nov 2020 to Apr 2021 | 81 | 40 (49.4%) | Ref |
|  | May 2021 to Oct 2021 | 171 | 83 (48.5%) | 0.99 (0.76, 1.29) |
|  | Nov 2021 to Apr 2022 | 86 | 26 (30.2%) | 0.73 (0.48, 1.11) |
|  | May 2022 to Oct 2022 | 93 | 27 (29.0%) | 0.68 (0.45, 1.02) |
|  | Nov 2022 to Apr 2023 | 40 | 18 (45.0%) | 1.07 (0.70, 1.65) |
|  |  | N=471 | ***Smoked more often than injected\**** |  |
|  | Nov 2020 to Apr 2021 | 81 | 7 (8.6%) | Ref |
|  | May 2021 to Oct 2021 | 171 | 17 (9.9%) | 1.14 (0.49, 2.64) |
|  | Nov 2021 to Apr 2022 | 86 | 14 (16.3%) | 2.69 (1.11, 6.49) |
|  | May 2022 to Oct 2022 | 93 | 15 (16.1%) | 2.58 (1.08, 6.16) |
|  | Nov 2022 to Apr 2023 | 40 | 9 (22.5%) | 3.49 (1.39, 8.74) |
| ***Fentanyl*** |  | N=480 | ***Injected\**** |  |
|  | Nov 2020 to Apr 2021 | 42 | 25 (59.5%) | Ref |
|  | May 2021 to Oct 2021 | 116 | 77 (66.4%) | 1.12 (0.85, 1.49) |
|  | Nov 2021 to Apr 2022 | 91 | 41 (45.1%) | 0.62 (0.43, 0.89) |
|  | May 2022 to Oct 2022 | 127 | 62 (48.8%) | 0.69 (0.50, 0.97) |
|  | Nov 2022 to Apr 2023 | 104 | 22 (21.2%) | 0.29 (0.18, 0.46) |
|  |  | N=480 | ***Smoked\**** |  |
|  | Nov 2020 to Apr 2021 | 42 | 35 (83.3%) | Ref |
|  | May 2021 to Oct 2021 | 116 | 98 (84.5%) | 1.02 (0.88, 1.20) |
|  | Nov 2021 to Apr 2022 | 91 | 77 (84.6%) | 1.08 (0.92, 1.26) |
|  | May 2022 to Oct 2022 | 127 | 112 (88.2%) | 1.12 (0.96, 1.31) |
|  | Nov 2022 to Apr 2023 | 104 | 102 (98.1%) | 1.26 (1.09, 1.46) |
|  |  | N=480 | ***Smoked more often than injected\**** |  |
|  | Nov 2020 to Apr 2021 | 42 | 25 (59.5%) | Ref |
|  | May 2021 to Oct 2021 | 116 | 70 (60.3%) | 1.03 (0.77, 1.37) |
|  | Nov 2021 to Apr 2022 | 91 | 64 (70.3%) | 1.29 (0.97, 1.72) |
|  | May 2022 to Oct 2022 | 127 | 81 (63.8%) | 1.16 (0.88, 1.54) |
|  | Nov 2022 to Apr 2023 | 104 | 101 (97.1%) | 1.80 (1.39, 2.33) |
| ***Opioids<sup>c</sup>*** |  | N=689 | ***Injected\**** |  |
|  | Nov 2020 to Apr 2021 | 84 | 83 (98.8%) | Ref |
|  | May 2021 to Oct 2021 | 186 | 174 (93.6%) | 0.95 (0.91, 1.00) |
|  | Nov 2021 to Apr 2022 | 133 | 97 (72.9%) | 0.69 (0.61, 0.78) |
|  | May 2022 to Oct 2022 | 164 | 115 (70.1%) | 0.67 (0.60, 0.76) |
|  | Nov 2022 to Apr 2023 | 122 | 53 (43.3%) | 0.41 (0.33, 0.51) |
|  |  | N=689 | ***Smoked\**** |  |
|  | Nov 2020 to Apr 2021 | 84 | 47 (56.0%) | Ref |
|  | May 2021 to Oct 2021 | 186 | 124 (66.7%) | 1.19 (0.96, 1.47) |
|  | Nov 2021 to Apr 2022 | 133 | 84 (63.2%) | 1.25 (1.00, 1.56) |
|  | May 2022 to Oct 2022 | 164 | 119 (72.6%) | 1.39 (1.13, 1.72) |
|  | Nov 2022 to Apr 2023 | 122 | 106 (86.9%) | 1.69 (1.37, 2.07) |
|  |  | N=689 | ***Smoked more often than injected\**** |  |
|  | Nov 2020 to Apr 2021 | 84 | 15 (17.9%) | Ref |
|  | May 2021 to Oct 2021 | 186 | 42 (22.6%) | 1.26 (0.75, 2.13) |
|  | Nov 2021 to Apr 2022 | 133 | 65 (48.9%) | 3.02 (1.86, 4.92) |
|  | May 2022 to Oct 2022 | 164 | 77 (47.0%) | 2.83 (1.74, 4.59) |
|  | Nov 2022 to Apr 2023 | 122 | 100 (82.0%) | 4.99 (3.13, 7.96) |
| ***Methamphetamine*** |  | N=719 | ***Injected\**** |  |
|  | Nov 2020 to Apr 2021 | 85 | 68 (80.0%) | Ref |
|  | May 2021 to Oct 2021 | 188 | 133 (70.7%) | 0.89 (0.77, 1.02) |
|  | Nov 2021 to Apr 2022 | 137 | 95 (69.3%) | 0.77 (0.64, 0.91) |
|  | May 2022 to Oct 2022 | 185 | 125 (67.6%) | 0.76 (0.64, 0.89) |
|  | Nov 2022 to Apr 2023 | 124 | 72 (58.1%) | 0.63 (0.52, 0.77) |
|  |  | N=719 | ***Smoked\**** |  |
|  | Nov 2020 to Apr 2021 | 85 | 78 (91.8%) | Ref |
|  | May 2021 to Oct 2021 | 188 | 170 (90.4%) | 0.98 (0.91, 1.07) |
|  | Nov 2021 to Apr 2022 | 137 | 111 (81.0%) | 0.87 (0.78, 0.97) |
|  | May 2022 to Oct 2022 | 185 | 161 (87.0%) | 0.94 (0.86, 1.02) |
|  | Nov 2022 to Apr 2023 | 124 | 121 (97.6%) | 1.05 (0.97, 1.13) |
|  |  | N=719 | ***Smoked more often than injected\**** |  |
|  | Nov 2020 to Apr 2021 | 85 | 38 (44.7%) | Ref |
|  | May 2021 to Oct 2021 | 188 | 112 (59.6%) | 1.34 (1.03, 1.74) |
|  | Nov 2021 to Apr 2022 | 137 | 80 (58.4%) | 1.43 (1.08, 1.89) |
|  | May 2022 to Oct 2022 | 185 | 123 (66.5%) | 1.61 (1.24, 2.08) |
|  | Nov 2022 to Apr 2023 | 124 | 109 (87.9%) | 2.18 (1.70, 2.79) |

<sup>a</sup> Obs = number of observations within each period, which includes only those who used each specified drug (i.e., heroin, fentanyl, opioids, methamphetamine) in the past six months (study participants within each period are unique); period one (November 2020-April 2021) includes only baseline observations, but the later periods include observations from multiple visits.
<sup>b</sup> Adjusted Relative Risk (aRR) for recruitment wave, age, gender, and recent (within the past six months) substance use treatment program enrollment.
<sup>c</sup> Represents use of heroin and/or fentanyl in the past six months.
\* Affirmative response, past six months.

Among participants who used fentanyl, risk of injecting it decreased by 71% from period one to period five (aRR=0.29, 95% CI 0.18, 0.46). Meanwhile, prevalence of smoking fentanyl remained high and stable between periods one and four, with risk of smoking fentanyl increasing by 26% in period five (vs. period one; aRR=1.26; 95% CI: 1.09, 1.46). In period five, risk of smoking fentanyl more often than injecting it increased by 80% (vs. period one; aRR=1.80, 95% CI 1.39, 2.33).

Among participants who used any opioids, risk of injecting them decreased by 59% from period one to period five (aRR=0.41; 95% CI: 0.33, 0.51). Risk of smoking opioids, meanwhile, increased substantially from period one, increasing in risk by 25% in period three (aRR=1.25; 95% CI: 1.00, 1.56), 39% in period four (aRR=1.39; 95% CI: 1.13, 1.72), and 69% in period five (aRR=1.69; 95% CI: 1.37, 2.07). Risk of smoking more often than injecting opioids also increased in periods three (aRR=3.02; 95% CI: 1.86, 4.92), four (aRR=2.83, 95% CI: 1.74, 5.49) and five (aRR=4.99; 95% CI: 3.13, 7.96) relative to period one.

Among participants who used methamphetamine, risk of injecting it declined by 23% in period three (aRR=0.77; 95% CI: 0.64, 0.91), 24% in period four (aRR=0.76; 95% CI: 0.64, 0.89) and 48% in period five (aRR=0.52; 95% CI: 0.52, 0.77) compared to period one. Risk of smoking methamphetamine declined by 13% in period three (vs. period one; aRR=0.87; 95% CI: 0.78, 0.97) but returned to period one levels in periods four and five. Risk of smoking methamphetamine more often than injecting it increased by 34% in period two (aRR=1.34; 95% CI: 1.03, 1.74), 43% in period three (aRR=1.43; 95% CI: 1.08, 1.89), 61% in period four (aRR=1.61; 95% 1.24, 2.08), and 118% in period five (aRR=2.18; 95% CI: 1.70, 2.79) relative to period one.

## 4. Discussion

This novel longitudinal assessment of drug consumption patterns among a cohort of people who injected drugs at baseline identified significant declines in the injection of opioids and methamphetamine from November 2020 to April 2023. We also observed marked increases in the prevalence of smoking these drugs and the likelihood of smoking these drugs more often than injecting them. These findings provide additional nuance on changing drug consumption patterns on the West Coast of the U.S. and have potential implications for future research and public health programming for people who use drugs. These revelations may be especially important in the context of rising overdose deaths driven by smoking drug use and polysubstance use with opioids and stimulants nationally (3, 7, 27).

Our study expands upon national and regional literature noting the diminishing prevalence of heroin use due to fentanyl’s infiltration into the drug supply and highlights a pronounced reduction in heroin use over time (23, 28, 29). We found a marked decline in both heroin injection and smoking in the overall sample, coupled with substantial increases in the risk of smoking it more often than injecting it over time among those recently using heroin.

In contrast, consistent with recently reported findings from San Francisco (14), fentanyl use patterns were more complex. Fentanyl injection, for instance, remained somewhat stable before decreasing significantly among those recently reporting its use. Meanwhile, fentanyl smoking increased overall, with a growing inclination towards smoking it more often than injecting it among those reporting its use. The observed increases in opioid smoking behaviors were likely primarily influenced by the rapid replacement of heroin with fentanyl in the U.S.-Mexico border region during our study period (23). However, a lack of consistently available drug-checking services in San Diego for most of our study period may have led some participants to misclassify heroin (or fentanyl) use when, in reality, they were using something else (30). The latter might further explain the shift from injecting to smoking in the context of heroin use. Altogether, our results imply an ongoing trend where injection of opioids is being supplanted by smoking.

This study examined previously unexplored patterns of methamphetamine consumption behaviors that extend our understanding of evolving polysubstance use trends. Methamphetamine injection declined by 50% over the study period and prevalence of smoking methamphetamine also declined overall (though less so than injecting). This could reflect behaviors amongst a fixed cohort rather than a population-level trend. Notably, smoking methamphetamine remained a common behavior in our sample and the overall pattern of transitioning from injecting to smoking was especially prominent for participants who used methamphetamine, where we saw a 100% increase in smoking methamphetamine more often than injecting it by the end of the study period. This trend may reflect the growing prevalence of polysubstance use involving methamphetamine and fentanyl (31–33), whereby methamphetamine may be used in combination to mitigate the heightened sedative effects of fentanyl, better cope with a more rapid onset of withdrawal symptoms and improve euphoric effects (34, 35). Also, the overall increase in smoking could potentially be due to the rising potency of illicitly manufactured fentanyls and methamphetamines that can create a strong effect that was not previously possible with heroin alone (34). Additional research that explicitly examines preferences in the co-administration of opioids and methamphetamine and why (and how) these two substances are so often used together is necessary.

The decreases in injecting and increases in smoking that we observed across these three unregulated drugs that are commonly used together may reflect changing community norms and preferences as well as prevention efforts focused on blood-borne infectious diseases (e.g., HIV, Hepatitis C, skin and soft tissue infections) and vein health (14, 15, 36, 37). Research is urgently needed to identify and quantify the positive and negative health impacts of smoking compared to injecting these drugs, as long-term risks may offset short-term benefits. While reduced transmission of blood-borne infectious diseases is a logical outcome of transitioning from injecting to smoking, the precise impacts of these transitions on overdose rates remain largely unknown, and possible harms to cardiovascular and lung health should be studied (38). Without such evidence, widespread beliefs that smoking (versus injecting) is “safer” could produce a sense of complacency and fewer harm-reducing behaviors (e.g., not carrying naloxone, increasing use while alone). Furthermore, the frequency of injections versus inhalation of fentanyl and methamphetamine—and related impacts of the length of time to onset of withdrawal symptoms among dependent individuals—deserves careful study. Indeed, it is possible that transitions to smoking lead to more frequent use and, hence, an increased risk of overdose. Ultimately, intervention and implementation research should aim to investigate strategies that promote equitable access to safer smoking supplies paired with harm reduction education, which could help connect people who smoke drugs to necessary services (27, 39).

In interpreting the results of this study, we acknowledge several limitations. First, reliance on self-reported behaviors introduces potential for misclassification, such as fentanyl being erroneously classified as heroin. Nevertheless, even in the presence of potential misclassification, the overall trends in injection remained unaffected, lending support to our interpretations. Second, trends for opioids (heroin and/or fentanyl) should be interpreted with caution since declines in opioid use are likely driven by decreased heroin use, while the increase in opioid smoking (and decrease in opioid injecting) is likely driven by changing fentanyl consumption behaviors. These differences indicate the importance of examining heroin and fentanyl use separately. Third, while efforts were made to minimize retention bias, the study experienced some loss to follow-up. Although we found minimal differences in baseline sociodemographics and substance use behaviors between those who completed at least one follow-up visit and those who did not, caution should be exercised in extrapolating these results to broader populations. However, the early impacts of drug trafficking seen in this region, such as changes to drug consumption trends, may nonetheless offer insights applicable to other areas, such as the West Coast of the U.S.

### 4.1. Conclusions

This longitudinal assessment of people who inject drugs from San Diego, California, revealed significant declines in the injection of heroin, fentanyl, and methamphetamine between November 2020 and April 2023. Notably, a substantial proportion of participants transitioned from injecting and smoking to only smoking by the end of the study period, regardless of drug classification. Also, the risk of smoking opioids more often than injecting increased over time, suggesting shifts in preference towards smoking among people using opioids. We also documented a marked increase in smoking methamphetamine more often than injecting it by the end of the study period. Taken together, these findings extend previous research identifying decreased injection and increased smoking of opioids by adding additional nuance concerning specific polysubstance use patterns. Further in-depth qualitative and quantitative research is needed to understand the determinants of these trends and their implications for the health of substance-using populations.

## Disclosures

### Role of funding source

This work was supported by the San Diego Center for AIDS Research (National Institute of Allergy and Infectious Diseases, grant P30AI036214) and the National Institute on Drug Abuse (grants R01DA049644-S1, R01DA049644-02S2, K01DA043412, 3K01DA043412-04S1, DP2DA049295, and T32DA023356). JRF was supported by the National Institute of General Medical Sciences (grant GM008042). The funders had no role in the decision to write and publish this manuscript or in the interpretation and presentation of findings.

### Contributors

Funding acquisition: S.A.S. and A.B.; Conceptualization, Methodology: W.H.E., S.A.S., D.A.; Data validation, curation, and analysis: D.A.; Original manuscript draft: W.H.E., A.R.B. and S.A.S.; Resources and project administration: C.F.V., A.H.V. and S.A.S. All authors contributed to the interpretation of results, manuscript revisions, and approved the final manuscript.

### Declarations of Interest

None.

### Conflict of Interest

We have no conflicts of interest to disclose.

## Acknowledgments

The authors gratefully acknowledge the *La Frontera* study team and participants.

## Data Availability Statement

Data is available upon reasonable request to the Principal Investigator of *La Frontera*, Dr. Steffanie Strathdee.

## Ethics Statement

Institutional review boards at UCSD and Xochicalco University approved all study activities.

## Notes

### Competing Interest Statement

The authors have declared no competing interest.

### Author Declarations

Institutional review boards at UCSD and Xochicalco University approved all study activities.

