## Supplemental tables for "Changes in injecting versus smoking heroin, fentanyl, and methamphetamine among people who inject drugs in San Diego, California, 2020 to 2023"

**SUPPLEMENTAL FILES**

**Supplemental Table 1. Survey window for each study visit for *La Frontera* by Cohort for study participants residing in San Diego County, California, between October 28, 2020, and April 27, 2023**

| **Dates** | **Visit Number** |
| --- | --- |
| **Cohort 1** |  |
| 10/28/2020-10/25/2021 | 1 |
| 05/07/2021-05/18/2022 | 2 |
| 12/07/2021-12/08/2022 | 3 |
| 06/03/2022-04/27/2023 | 4 |
| 11/22/2022-04/24/2023 | 5 |
| **Cohort 2** |  |
| 02/07/2022-06/09/2022 | 1 |
| 08/24/2022-03/21/2023 | 2 |
| 01/04/2023-04/27/2023 | 3 |

**Supplemental Table 2. Prevalence of only injecting, only smoking, and injecting and smoking (heroin, fentanyl, or methamphetamine) in the past six months among study participants residing in San Diego County, California, between October 28, 2020, and April 31, 2023**

| **Time Period** | **Obs^a^** | **Only injected^b^**  **N (%)** | **Only smoked^b^**  **N (%)** | **Injected and smoked^b^**  **N (%)** |
| --- | --- | --- | --- | --- |
|  | N=833 |  |  |  |
| Nov 2020 to Apr 2021 | 96 | 15 (15.6%) | 0 (0.0%) | 81 (84.4%) |
| May 2021 to Oct 2021 | 212 | 26 (12.3%) | 7 (3.3%) | 179 (84.4%) |
| Nov 2021 to Apr 2022 | 167 | 32 (19.2%) | 21 (12.6%) | 114 (68.3%) |
| May 2022 to Oct 2022 | 203 | 25 (12.3%) | 39 (19.2%) | 139 (68.5%) |
| Nov 2022 to Apr 2023 | 155 | 10 (6.5%) | 53 (34.2%) | 92 (59.4%) |

^a^ Obs = number of observations (study participants within each period are unique; period one (November 2020-April 2021) includes only baseline observations, but the later periods include observations from multiple visits).

^b^ Mode of consumption for heroin, fentanyl, or methamphetamine in the past six months.

Abbreviations: Nov = November; Apr = April; Oct = October.
